# Clinical Outcomes From COVID-19 Following Use of Angiotensin-Converting Enzyme Inhibitors or Angiotensin-Receptor Blockers Among Patients with Hypertension in South Korea: A nationwide study

**DOI:** 10.1101/2020.07.29.20164822

**Authors:** Ju Hwan Kim, Yeon-Hee Baek, Hyesung Lee, Young June Choe, Hyun Joon Shin, Ju-Young Shin

## Abstract

There is ongoing debate as to whether angiotensin-converting enzyme inhibitors (ACEIs) or angiotensin-receptor blockers (ARBs) use is associated with poor prognosis of coronavirus disease-2019 (COVID-19). We sought to investigate the association between ACEI/ARB use and risk of poor clinical outcomes from COVID-19. We identified 1,290 patients with hypertension, of which 682 had recorded ACEI/ARB use and 608 without the use during 30 days preceding the date of COVID-19 diagnosis in completely enumerated COVID-19 cohort in South Korea. Our primary endpoint was the clinical outcomes comprised of all-cause mortality, use of mechanical ventilation, intensive care unit (ICU) admission, and sepsis. We used inverse probability of treatment weighting (IPTW) to mitigate selection bias, and Poisson regression model to estimate the relative risks (RR) and 95% confidence intervals (CI) to compare outcomes in ACEI/ARB users with non-users. Compared to non-use, ACEI/ARB use was associated with lower clinical outcomes (IPTW adjusted RR, 0.60; 95% CI, 0.42-0.85; p=0.0046). When assessed by individual outcomes, ACEI/ARB use was not associated with all-cause mortality (IPTW adjusted RR, 0.62; 95% CI, 0.35-1.09; p=0.0973) and respiratory events (IPTW adjusted RR, 0.99; 95% CI, 0.84-1.17; p=0.9043). Subgroup analysis showed a trend toward protective role of ACEIs and ARBs against overall outcomes in men (IPTW adjusted RR, 0.84; 95% CI, 0.69-1.03; p-for-interaction=0.008) and with pre-existing respiratory disease (IPTW adjusted RR, 0.74; 95% CI, 0.60-0.92; p-for-interaction=0.0023). We present clinical evidence to support continuing ACE/ARB use in completely enumerated hypertensive COVID-19 cohort in South Korea.

## Introduction

The rapid spread of coronavirus disease 2019 (COVID-19), caused by severe acute respiratory syndrome coronavirus 2 (SARS-CoV-2), is an ongoing pandemic, having infected >2,800,000 people over 213 countries as of April 28, 2020 ^1^. Concerns have been raised regarding poor prognosis of COVID-19 associated with the use of angiotensin-converting enzyme inhibitors (ACEI) and angiotensin II receptor blockers (ARBs) ^2^. ACEIs and ARBs were shown to upregulate ACE2 expression and activity in several experimental studies ^3-5^. Given that the binding of ACE2 and viral spike protein of SARS-CoV-2 allows coronavirus entry into host cells, it had been hypothesized that potential upregulation of ACE2 owing to use of ARBs leads to an increased risk of severe and fatal health outcomes ^2,6^.

In theory, patients with hypertension being managed with ACEIs or ARBs could be at increased risk of COVID-19 since increased ACE2 by these drug classes may increase viral entry into cells. Alternatively, increased ACE2 had been recognized to counterbalance the pro-inflammatory and vasoconstrictive effect of ACE, mainly through conversion of angiotensin II (Ang II) to Ang-(1-7), a peptide with potential protective anti-inflammatory properties that counterbalances pro-inflammatory activity of Ang II ^7-9^.

In view of these two opposing mechanistic hypotheses, demands for clinical research remain very high. To date, several observational studies claimed that the use of ACEI/ARB was not associated with increased all-cause mortality ^10-14^. However, these studies were limited due to methodological issues in study design, with the former lacking causal relationship assessment and latter suffering from immortal time bias arising from misclassification of exposure period. Given the lack of robust population-based study assessing the association between the use of ACEI/ARB and sequelae of COVID-19, we analyzed completely enumerated hypertensive COVID-19 cohort in South Korea to assess whether the use of ACEIs and ARBs are associated with poor clinical outcomes from COVID-19.

## Methods

We retrieved healthcare database from the Health Insurance Review & Assessment Service of South Korea, which covers over 50 million entire South Korean population, from January 1, 2015 to April 8, 2020. We used completely enumerated database of 69,793 subjects who underwent COVID-19 screening test in South Korea. The database contains both inpatient and outpatient prescriptions, demographic (age, sex, and insurance type) and clinical information on visit dates for hospitalization and ambulatory cares, procedures, and diagnosis records coded using Korea Standard Classification of Diseases, 7^th^ revision (K CD-7), which is based on International Classification of Diseases, 10^th^ revision (ICD-10). The overall agreement of diagnostic records of hypertension, stroke, and heart disease was 93.73%, 98.80%, and 97.93%, respectively ^15^.

We conducted a retrospective cohort study of ACEI/ARB use and adverse outcomes from COVID-19 among patients with hypertension. We identified patients with laboratory confirmed diagnosis of COVID-19 between December 1, 2019 and April 8, 2020. Diagnoses were made based on the diagnostic results from on reverse transcription polymerase chain reaction (RT-PCR) method targeting the RNA-dependent RNA polymerase (RdRP), N, and E genes as recommended by the interim guidance of World Health Organization ^16^. Cohort entry was defined as the date of incident diagnosis of COVID-19. We required patients to have a recorded diagnosis of hypertension within a 5-year lookback period from the cohort entry.

Exposure to ACEIs and ARBs was ascertained within 30 days preceding the cohort entry. Our exposure of interest was patients ever prescribed with ACEIs or ARBs either as a monotherapy or combination therapy. Non-users were those who had no prescription record of either ACEIs or ARBs during the exposure ascertainment period.

Our study outcomes included clinical outcomes indicative of poor prognosis for COVID-19, comprised of all-cause mortality, use of mechanical ventilation, intensive care unit (ICU) admission, or sepsis. We also assessed all-cause mortality and respiratory events (acute respiratory distress syndrome, interstitial lung disease, pneumonia, and respiratory failure) individually as the secondary endpoints. Each patient was followed until the occurrence of outcome of interest or data-censoring date.

We assessed baseline characteristics within 1 year before cohort entry. To make propensity score we used baseline confounders which include age at cohort entry, sex, income level, CHA□DS□-VASc score, medical history (including diabetes, cardiovascular disease (CVD), stroke, other cerebrovascular disease, hyperlipidemia, respiratory disease, chronic kidney disease, cancer, thromboembolism, and dementia), comedications (including other antihypertensives (calcium channel blockers (CCBs), diuretics, β-blockers, and alpha blockers), antidiabetics, antibiotics, antiarrhythmics, antiplatelets, anticoagulants, lipid-lowering agents, and antianginal agents), dialysis, and the duration of hypertension (<1 year, ≥1 year and <3 years, ≥3 years and <5 years, and ≥5 years). The treatments for COVID-19 (including antibiotics, antivirals, antimalarials (chloroquine and hydroxychloroquine), corticosteroids, and intravenous immunoglobulin) were assessed but not included in the propensity score model as they act as the intermediates between exposure and outcome of our study.

We estimated propensity scores for receiving ACEIs or ARBs by fitting a multivariable logistic regression model using all pre-defined covariates assessed 1 year before cohort entry. We used inverse probability of treatment weighting (IPTW) based on the propensity scores to mitigate selection bias by the characteristics between ACEI/ARB users and non-users. IPTW creates a pseudo population, where the weighted exposure and comparator groups are representative of the patient characteristics in the overall population, and thus generates the population average treatment effect ^17^. We summarized baseline characteristics of the study cohort using counts and proportions or mean for categorical or continuous variables, respectively. We used Poisson regression model to estimate relative risk (RR) and corresponding 95% confidence intervals (CI) of each outcome in ACEI/ARB users compared to non-users in patients with COVID-19. The unweighted model was adjusted for pre-defined covariates including age, sex, CHA□DS□-VASc score, diabetes, CVD, and baseline respiratory diseases for parsimony. These covariates were also used in the IPTW weighted models for doubly robust estimation of causal effect. We chose adjusted IPTW weighted model as a main model to report the RRs and corresponding 95% CIs for the clinical outcomes in ACEI/ARB users, compared with non-user.

Given that ACEIs and ARBs, apart from hypertension, are primarily prescribed for patients with diabetes and CVD, we repeated analysis with the restricted cohort of patients with these health conditions to exclude confounding by indication. To evaluate whether the association differed by underlying patients’ conditions, we conducted additional subgroup analysis using the interaction terms by age group, sex, CHA□DS□-VASc score, pre-existing respiratory disease, and hospitalization after diagnosis of COVID-19. In subgroup analysis, we used overall outcomes which include all-cause mortality, use of mechanical ventilation, ICU admission, sepsis, or the occurrence of respiratory events to increase the statistical power. Additionally, we conducted sensitivity analysis where we redefined exposure assessment window as within 180 days preceding the cohort entry to address potential exposure misclassification.

Our study complies with the Declaration of Helsinki and the study protocol was approved by the Institutional Review Board (IRB) of Sungkyunkwan University (SKKU 2020-03-021) and obtaining informed consent was waived by the IRB.

## Results

Among 5,707 patients with confirmed diagnosis of COVID-19, there were 1,290 patients with past medical history of hypertension, of which 682 had recorded use of ACEI/ARB and 608 without the use during 30 days preceding cohort entry (**Figure 1**). The characteristics of the ACEI/ARB users compared with non-users are described in Table 1. Compared to non-users, ACEI/ARB users were older (mean age (standard deviation) 62.8 years (14.4) vs. 61.3 years (16.6)), had higher proportion of men (53.4% vs. 49.8%), higher prevalence of hyperlipidemia (38.6% vs. 33.6%), diabetes (37.0% vs. 25.7%), CVD (27.9% vs. 26.0%), chronic kidney disease (18.8% vs. 15.6%) and duration of hypertension over 5 years (56.7% vs. 41.5%). Concomitant use of other anti-hypertensives was generally similar between ACEI/ARB users and non-users, while the mean CHA_2_DS_2_-VASc score was higher in ACEI/ARB users (2.7 vs. 2.4).

**Table 1.** Characteristics of ACEI/ARB users and non-users in hypertensive COVID-19 patients

| Characteristics | Overall Cohort (n=1,290) | ACEI/ARB use (n=682) | Non-use (n=608) |
| --- | --- | --- | --- |
| **Age years, mean (SD)** | 62.1 (15.5) | 62.8 (14.4) | 61.3 (16.6) |
| <40 | 114 (8.8) | 42 (6.2) | 72 (11.8) |
| 40-64 | 582 (45.1) | 325 (46.7) | 257 (42.3) |
| 65-84 | 517 (40.1) | 273 (40.0) | 244 (40.1) |
| ≥85 | 77 (6.0) | 42 (6.2) | 35 (5.8) |
| **Male, N (%)** | 667 (51.7) | 364 (53.4) | 303 (49.8) |
| **Medical History, N (%)** |  |  |  |
| Hyperlipidemia | 467 (36.2) | 263 (38.6) | 204 (33.6) |
| Diabetes | 408 (31.6) | 252 (37.0) | 156 (25.7) |
| Cancer | 138 (10.7) | 62 (9.1) | 76 (12.5) |
| **Respiratory disease** | 432 (33.5) | 222 (32.6) | 210 (34.5) |
| Asthma | 132 (10.2) | 59 (8.7) | 73 (12.0) |
| COPD | 283 (21.9) | 137 (20.1) | 146 (24.0) |
| Bronchiectasis | 15 (1.2) | 10 (1.5) | 5 (0.8) |
| Pneumonia | 143 (11.1) | 83 (12.2) | 60 (9.9) |
| Interstitial lung disease | 16 (1.24) | 8 (1.2) | 8 (1.3) |
| **Cardiovascular disease** | 348 (27.0) | 190 (27.9) | 158 (26.0) |
| Peripheral vascular disease | 113 (8.8) | 64 (9.4) | 49 (8.1) |
| Coronary artery disease | 150 (11.6) | 72 (10.6) | 78 (12.8) |
| Atrial fibrillation | 40 (3.1) | 19 (2.8) | 21 (3.5) |
| Valvular heart disease | 10 (0.8) | 8 (1.2) | 2 (0.3) |
| Heart failure | 100 (7.8) | 59 (8.7) | 41 (6.7) |
| Arrhythmia | 38 (3.0) | 18 (2.6) | 20 (3.3) |
| Chronic kidney disease | 223 (17.3) | 128 (18.8) | 95 (15.6) |
| Chronic liver disease | 164 (12.7) | 80 (11.7) | 84 (13.8) |
| Stroke | 137 (10.6) | 63 (9.2) | 74 (12.2) |
| Other cerebrovascular diseases | 87 (6.7) | 44 (6.5) | 43 (7.1) |
| **Comedications, N (%)** |  |  |  |
| CCBs | 476 (36.9) | 228 (33.4) | 248 (40.8) |
| Diuretics | 282 (21.9) | 149 (21.9) | 133 (21.9) |
| β-blockers | 368 (28.5) | 182 (26.7) | 186 (30.6) |
| Alpha blockers | 187 (14.5) | 106 (15.5) | 81 (13.3) |
| **CHA<sub>2</sub>DS<sub>2</sub>-VASc score, mean (SD)** | 2.5 (1.6) | 2.7 (1.4) | 2.4 (1.7) |
| 0-1 | 371 (28.8) | 148 (21.7) | 223 (36.7) |
| 2-5 | 860 (66.7) | 507 (74.3) | 353 (58.1) |
| 6-9 | 59 (4.6) | 27 (4.0) | 32 (5.3) |
| **Duration of hypertension, N (%)** |  |  |  |
| <1 year | 114 (8.8) | 40 (5.9) | 74 (12.2) |
| ≥1 year and <3 years | 247 (19.2) | 106 (15.5) | 141 (23.2) |
| ≥3 years and <5 years | 290 (22.5) | 149 (21.9) | 141 (23.2) |
| ≥5 years | 639 (49.5) | 387 (56.7) | 252 (41.5) |

Abbreviations: ACEIs, angiotensin-converting enzyme inhibitors; ARBs, angiotensin II receptor blockers; COVID-19, coronavirus disease 2019; SD, standard deviation; COPD, chronic obstructive pulmonary disease; CCBs, calcium channel blockers.

**Figure 1.**
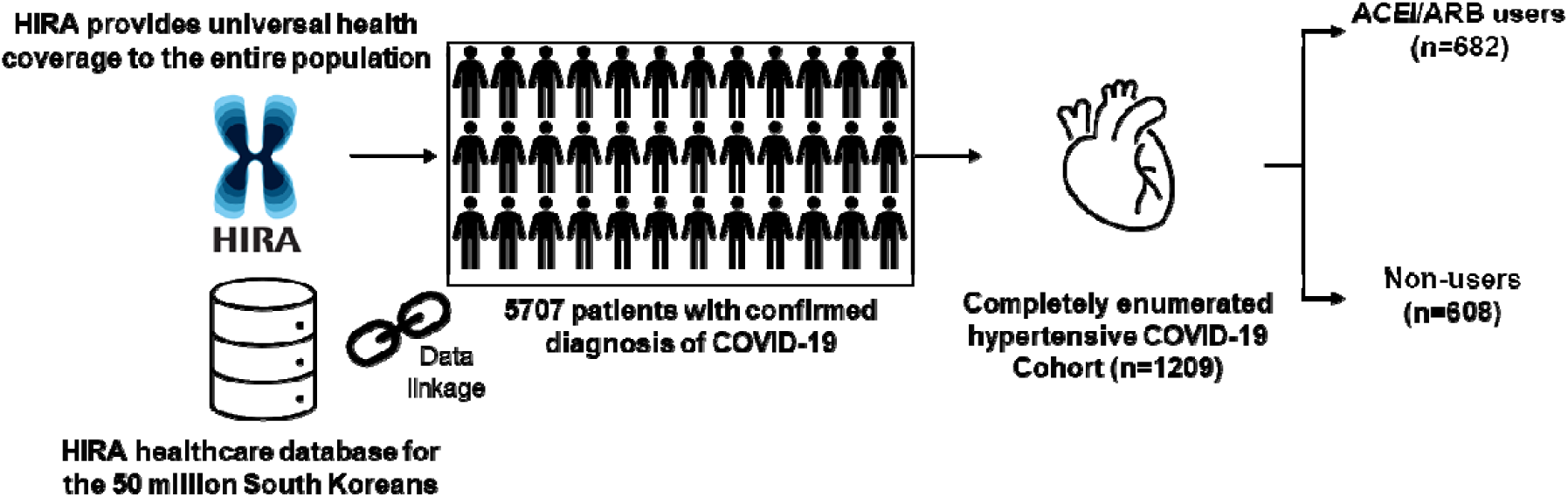
Description of data source and selection of hypertensive COVID-19 cohort. Abbreviations: HIRA, Health Insurance Review & Assessment Service; COVID-19, coronavirus disease 2019; ACEI, angiotensin converting enzyme inhibitor; ARB, angiotensin II receptor blocker.

During the study period, there were 23 (3.4%) and 28 (4.6%) cases of clinical outcomes in ACEI/ARB users and non-users, respectively (**Table 2**). Compared to non-use, ACEI/ARB use was associated with lower clinical outcomes that included all-cause mortality, mechanical ventilation, ICU admission, and sepsis (IPTW adjusted RR, 0.60; 95% CI, 0.42-0.85; p=0.0046). When assessed by individual outcome event, ACEI/ARB use was not associated with the risk of all-cause mortality (IPTW adjusted RR, 0.62; 95% CI, 0.35-1.09; p=0.0973) and respiratory events (IPTW adjusted RR, 0.99; 95% CI, 0.84-1.17; p=0.9043) compared with non-use.

**Table 2.**
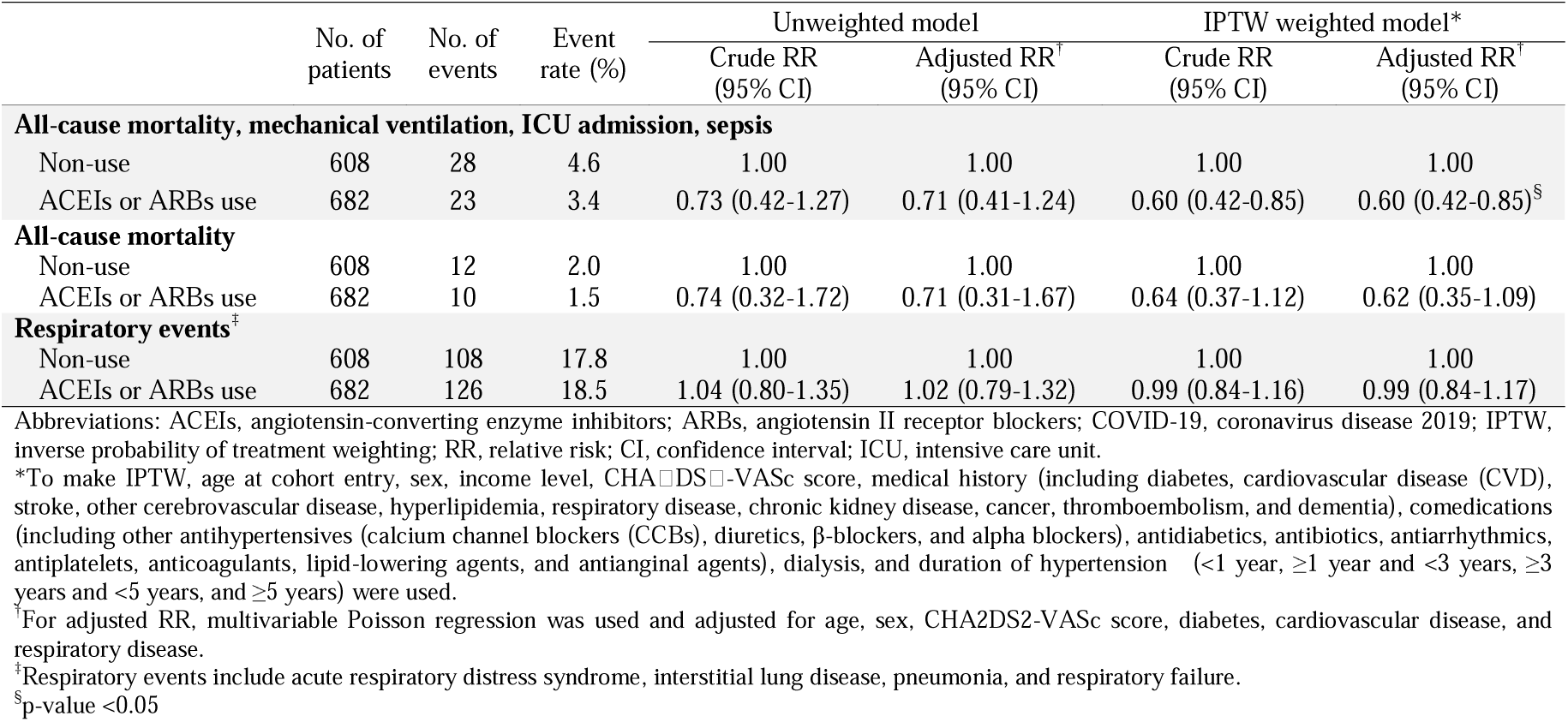
Relative risks of clinical outcomes in ACEI/ARB users compared to non-users in in hypertensive COVID-19 patients

|  | No. of patients | No. of events | Event rate (%) | Unweighted model |  | IPTW weighted model* |  |
| --- | --- | --- | --- | --- | --- | --- | --- |
|  |  |  |  | Crude RR (95% CI) | Adjusted RR<sup>†</sup> (95% CI) | Crude RR (95% CI) | Adjusted RR<sup>†</sup> (95% CI) |
| All-cause mortality, mechanical ventilation, ICU admission, sepsis |  |  |  |  |  |  |  |
| Non-use | 608 | 28 | 4.6 | 1.00 | 1.00 | 1.00 | 1.00 |
| ACEIs or ARBs use | 682 | 23 | 3.4 | 0.73 (0.42-1.27) | 0.71 (0.41-1.24) | 0.60 (0.42-0.85) | 0.60 (0.42-0.85)<sup>§</sup> |
| All-cause mortality |  |  |  |  |  |  |  |
| Non-use | 608 | 12 | 2.0 | 1.00 | 1.00 | 1.00 | 1.00 |
| ACEIs or ARBs use | 682 | 10 | 1.5 | 0.74 (0.32-1.72) | 0.71 (0.31-1.67) | 0.64 (0.37-1.12) | 0.62 (0.35-1.09) |
| Respiratory events<sup>‡</sup> |  |  |  |  |  |  |  |
| Non-use | 608 | 108 | 17.8 | 1.00 | 1.00 | 1.00 | 1.00 |
| ACEIs or ARBs use | 682 | 126 | 18.5 | 1.04 (0.80-1.35) | 1.02 (0.79-1.32) | 0.99 (0.84-1.16) | 0.99 (0.84-1.17) |

Abbreviations: ACEIs, angiotensin-converting enzyme inhibitors; ARBs, angiotensin II receptor blockers; COVID-19, coronavirus disease 2019; IPTW, inverse probability of treatment weighting; RR, relative risk; CI, confidence interval; ICU, intensive care unit.
\*To make IPTW, age at cohort entry, sex, income level, CHA<sub>2</sub>DS<sub>2</sub>-VASc score, medical history (including diabetes, cardiovascular disease (CVD), stroke, other cerebrovascular disease, hyperlipidemia, respiratory disease, chronic kidney disease, cancer, thromboembolism, and dementia), comedications (including other antihypertensives (calcium channel blockers (CCBs), diuretics, β-blockers, and alpha blockers), antidiabetics, antibiotics, antiarrhythmics, antiplatelets, anticoagulants, lipid-lowering agents, and antianginal agents), dialysis, and duration of hypertension (<1 year, $\geq 1$ year and <3 years, $\geq 3$ years and <5 years, and $\geq 5$ years) were used.
<sup>†</sup>For adjusted RR, multivariable Poisson regression was used and adjusted for age, sex, CHA<sub>2</sub>DS<sub>2</sub>-VASc score, diabetes, cardiovascular disease, and respiratory disease.
<sup>‡</sup>Respiratory events include acute respiratory distress syndrome, interstitial lung disease, pneumonia, and respiratory failure.
<sup>§</sup>p-value <0.05

We conducted sensitivity analysis where we redefined the exposure risk window as 180 days preceding the cohort entry to account for potential exposure misclassification (**Table 3**). There were 28 (3.7%) and 23 (4.4%) cases of adverse outcomes in ACEI/ARB users and non-users, respectively. The results from sensitivity analysis were generally consistent with the main analysis; ACEI/ARB use was associated with lower clinical outcomes (IPTW adjusted RR, 0.65; 95% CI, 0.46-0.90; p=0.0094), all-cause mortality (IPTW adjusted RR, 0.41; 95% CI, 0.25-0.68; p=0.0006), but was not associated with respiratory events (IPTW adjusted RR, 0.94; 95% CI, 0.81-1.09; p=0.4188) compared with non-use.

**Table 3.** Sensitivity analysis with redefined exposure risk window of 180 days preceding the cohort entry for the relative risks of clinical outcomes in ACEI/ARB users compared to non-users in hypertensive COVID-19 patients

|  | No. of patients | No. of events | Event rate (%) | Unweighted model |  | IPTW weighted model* |  |
| --- | --- | --- | --- | --- | --- | --- | --- |
|  |  |  |  | Crude RR (95% CI) | Adjusted RR<sup>†</sup> (95% CI) | Crude RR (95% CI) | Adjusted RR<sup>†</sup> (95% CI) |
| All-cause mortality, mechanical ventilation, ICU admission, sepsis |  |  |  |  |  |  |  |
| Non-use | 523 | 23 | 4.4 | 1.00 | 1.00 | 1.00 | 1.00 |
| ACEIs or ARBs use | 767 | 28 | 3.7 | 0.83 (0.48-1.44) | 0.79 (0.45-1.38) | 0.68 (0.49-0.94)<sup>§</sup> | 0.65 (0.46-0.90)<sup>§</sup> |
| All-cause mortality |  |  |  |  |  |  |  |
| Non-use | 523 | 12 | 2.3 | 1.00 | 1.00 | 1.00 | 1.00 |
| ACEIs or ARBs use | 767 | 10 | 1.3 | 0.57 (0.25-1.32) | 0.52 (0.22-1.22) | 0.45 (0.27-0.74)<sup>§</sup> | 0.41 (0.25-0.68)<sup>§</sup> |
| Respiratory events<sup>‡</sup> |  |  |  |  |  |  |  |
| Non-use | 523 | 92 | 17.6 | 1.00 | 1.00 | 1.00 | 1.00 |
| ACEIs or ARBs use | 767 | 142 | 18.5 | 1.05 (0.81-1.37) | 1.08 (0.77-1.32) | 0.98 (0.84-1.13) | 0.94 (0.81-1.09) |

Abbreviations: ACEIs, angiotensin-converting enzyme inhibitors; ARBs, angiotensin II receptor blockers; COVID-19, coronavirus disease 2019; IPTW, inverse probability of treatment weighting; RR, relative risk; CI, confidence interval; ICU, intensive care unit.
\*To make IPTW, age at cohort entry, sex, income level, CHA<sub>2</sub>DS<sub>2</sub>-VASc score, medical history (including diabetes, cardiovascular disease (CVD), stroke, other cerebrovascular disease, hyperlipidemia, respiratory disease, chronic kidney disease, cancer, thromboembolism, and dementia), comedications (including other antihypertensives (calcium channel blockers (CCBs), diuretics, β-blockers, and alpha blockers), antidiabetics, antibiotics, antiarrhythmics, antiplatelets, anticoagulants, lipid-lowering agents, and antianginal agents), dialysis, and duration of hypertension (<1 year, ≥ 1 year and <3 years, ≥ 3 years and <5 years, and ≥ 5 years) were used.
<sup>†</sup>For adjusted RR, multivariable Poisson regression was used and adjusted for age, sex, CHA<sub>2</sub>DS<sub>2</sub>-VASc score, diabetes, cardiovascular disease, and respiratory disease.
<sup>‡</sup>Respiratory events include acute respiratory distress syndrome, interstitial lung disease, pneumonia, and respiratory failure.
<sup>§</sup>p-value <0.05

Subgroup analysis on the risk of clinical outcomes compared with non-use is presented in **Figure 2**. When assessed by exposure subtypes, no significant interaction between the subtypes and the overall outcomes was found (p for interaction=0.015); ACEI (IPTW adjusted RR, 0.67; 95% CI, 0.42-1.06) and ARB use (IPTW adjusted RR, 0.97; 95% CI, 0.83-1.13) was not associated with the risk of overall adverse outcomes. Interestingly, interaction-term analysis showed a trend toward protective role of ACEIs and ARBs against overall outcomes in men (IPTW adjusted RR, 0.84; 95% CI, 0.69-1.03; p for interaction=0.008), with pre-existing respiratory disease (IPTW adjusted RR, 0.74; 95% CI, 0.60-0.92; p for interaction=0.0023) and in patients hospitalized for COVID-19 (IPTW adjusted RR, 0.93; 95% CI, 0.78-1.10; p for interaction<0.0001).

**Figure 2.**
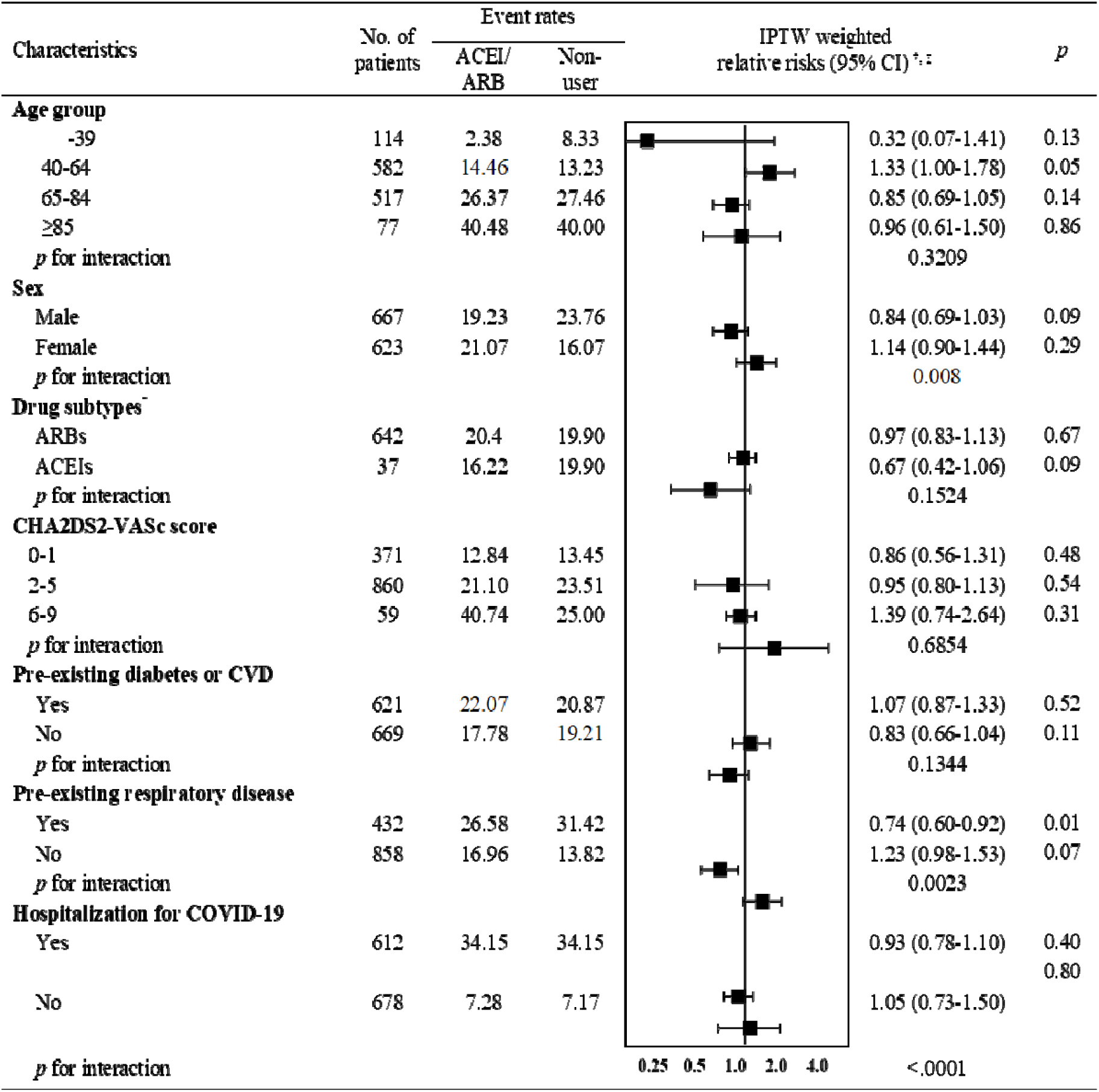
Relative risks (95% confidence intervals) of overall clinical outcomes* in ACEI/ARB users compared to non-users in selected population subgroups. Abbreviations: ACEIs, angiotensin-converting enzyme inhibitors; ARBs, angiotensin II receptor blockers; IPTW, inverse probability of treatment weighting; CI, confidence interval *Overall clinical outcomes include all-cause mortality, use of mechanical ventilation, admission to intensive care unit, sepsis, or the occurrence of respiratory events. ^†^ Relative risks were adjusted for age, sex, CHA2DS2-VASc score, diabetes, cardiovascular disease, and respiratory disease. ^‡^ To make IPTW, age at cohort entry, sex, income level, CHA□DS□-VASc score, medical history (including diabetes, cardiovascular disease (CVD), stroke, other cerebrovascular disease, hyperlipidemia, respiratory disease, chronic kidney disease, cancer, thromboembolism, and dementia), comedications (including other antihypertensives (calcium channel blockers (CCBs), diuretics, β-blockers, and alpha blockers), antidiabetics, antibiotics, antiarrhythmics, antiplatelets, anticoagulants, lipid-lowering agents, and antianginal agents), dialysis, and the duration of hypertension (<1 year, ≥1 year and <3 years, ≥3 years and <5 years, and ≥5 years).

## Discussion

To the best of our knowledge, this retrospective cohort study provides the first completely enumerated hypertensive COVID-19 cohort study in a national scale. We used medical claims data of patients diagnosed with COVID-19 in South Korea to demonstrate that ACEI/ARB use was not associated with poor clinical outcomes from COVID-19 among patients with hypertension. Specifically, ACEI/ARB use, compared with non-use, was associated with lower clinical outcomes that comprised of all-cause mortality, use of mechanical ventilation, ICU admission, and sepsis. To account for exposure misclassification, we conducted sensitivity analysis to assess exposure status during a period of 180 days preceding the cohort entry in which the IPTW adjusted RRs were largely consistent with the findings from main analysis across all outcome measures. Furthermore, the results from subgroup analysis accounting for potential confounding by indication also remained largely consistent with the findings from main analysis.

While the underlying pathogenic link between hypertension and COVID-19 remains to be elucidated, concerns have been raised that ACEIs and ARBs, mainstay of therapy for hypertension and diabetes, may contribute to the adverse outcomes observed in COVID-19 patients ^2^. Indeed, interaction between SARS-CoV-2 and ACE2 was proposed a potential mechanism for COVID-19’s cell entry ^18^, and administration of ACEIs and ARBs upregulated ACE2 expression and activity in several experimental studies ^3-5^, which may theoretically predispose patients on ACEIs or ARBs at greater risk of COVID-19. Conversely, beneficial role of ACE2 in COVID-19 had been reported, of which a recent pilot clinical trial in acute respiratory distress syndrome demonstrated the promising role of recombinant human ACE2 in attenuating the acute lung injury ^19^. Moreover, experimental evidence indicates that ARBs, specifically losartan, restored the expression level of ACE2 which was downregulated in pre-clinical models of experimental SARS-CoV infection and acute lung injury ^3,20,21^. While there is an ongoing debate on whether to continue or halt ACEI/ARB in COVID-19 patients with hypertension, real-world data from our study complement the position statements made by the medical societies such as European Society of Cardiology council, American College of Cardiology, American Heart Association, and Heart Failure Society of America on continuing the use of ACEIs or ARBs as prescribed ^22,23^, as ACEI/ARB use was not associated with poor clinical outcomes from COVID-19.

Consistent with our study finding, several recently published studies also have demonstrated no harm or even protective role of ACEI/ARB in COVID-19. A study in Italy utilized case-control design that involved 6,272 COVID-19 cases and 30,579 matched controls to report that ACEI/ARB use was not associated with the risk of COVID-19 (adjusted OR, 0.95; 95% CI, 0.86-1.05) ^13^. The most recent single tertiary center-based study in the United States also reported null association between ACEI/ARB use and poor outcomes from COVID-19 among 2,573 hypertensive COVID-19 patients, with the median difference in percentage points between ACEI/ARB users and non-users of −0.5% (95% CI, - 4.3-3.2) ^14^. Although methodological issues inherent in observational studies limit the interpretation of the study findings, study conclusions of these recently published studies are consistent with the findings of our study, and provide clinical evidence that ACEI/ARB use, at least, are not associated with increased risk of poor clinical outcomes from COVID-19. In addition to the observed protective role of ACEIs and ARBs during COVID-19, our subgroup analysis showed a greater benefit with regard to clinical outcomes from COVID-19 in association with ACEI/ARB use than with non-use in men, with pre-existing respiratory disease, and in patients hospitalized for COVID-19. These subgroups had been reported to have poor prognosis in COVID-19 ^13^, and our study findings should be interpreted with caution as we used overall outcomes to increase the statistical power in assessing the role of ACEI/ARB among these subgroups. Additional aspect to be noted in our subgroup analysis is that proportion of patients taking ARBs were notably higher compared with ACEIs as regional hypertension management guideline in Korea recommended ARBs over ACEIs due to more favorable adherence and less frequent adverse events ^24^.

Our study provided clinical evidence indicating ACEI/ARB use not associated with poor prognosis of COVID-19. We generated practicable evidence which addresses the urgent public health need in the uncertainty of clinical consequences of ACEI/ARB use among patients with COVID-19. Second, our results have a solid external validity by assembling completely enumerated COVID-19 cases that occurred in South Korea. South Korea have implemented rigorous screening, contact tracing and quarantine measures, conducting a total of 601,660 COVID-19 screening tests as of April 27 to proactively contain COVID-19 ^25^. All individuals with epidemiologic links with suspected or confirmed COVID-19 patients or came from abroad have been self-quarantined for 14 days, and those who developed a fever (37.5°C and above) or respiratory symptoms received COVID-19 screening tests ^26^; thus, underdiagnosis of COVID-19 is likely to be minimal. Third, our study results were consistent in the subgroup analysis by pre-existing diabetes or CVD, which suggests the robustness of our results from confounding by indication given that ACEIs and ARBs are primarily prescribed for patients with these coexisting comorbidities.

Our study also has some limitations. First, potential misclassification of diagnosis-based outcomes (sepsis and respiratory events) may present. Nevertheless, a validation study comparing diagnoses in the South Korean healthcare database with electronic medical records reported an overall positive predictive value of 82% ^27^. Death records and procedure codes including mechanical ventilation and ICU admission have high validity, and thus less likely to affect our study conclusion. Second, there is a potential exposure misclassification owing to short exposure ascertainment period. We found consistent result with the main analysis when the exposure risk window was redefined as 180 days. Third, residual confounding from unmeasured confounders (e.g. smoking history, body mass index, baseline blood pressure and laboratory test results) may have affected our results given inherent limitation of available variables in the analysis of health claims data. Finally, we included prevalent users of ACEI/ARB, while ideally new-user design is recommended where all study subjects are naïve to previous use of ACEI/ARB to address potential under ascertainment of events that occur early in therapy and to precisely control for confounders that may be altered by the study drug ^28^. However, we used a prevalent user cohort of ACEI/ARB given that a new-user design would exclude the large number of subjects that may represent clinically relevant subset.

In conclusion, our study findings did not identify increased risk of adverse outcomes with the use of ACEIs or ARBs among COVID-19 patients with hypertension. We present clinical evidence to support current medical societies recommendation on continuing ACEIs or ARBs as prescribed in COVID-19 patients.

## Data Availability

No additional data available

https://hira-covid19.net/

## Acknowledgement

The authors appreciate healthcare professionals dedicated to treating COVID-19 patients in Korea, and the Ministry of Health and Welfare, the Health Insurance Review & Assessment Service, and Do-Yeon Cho of the Health Insurance Review & Assessment Service of South Korea for sharing invaluable national health insurance claims data in a prompt manner.

